# Elevating the patient perspective: Qualitative evaluation of non-U.S.–born care navigation on latent tuberculosis infection screening and treatment adherence

**DOI:** 10.64898/2026.06.04.26354954

**Authors:** Laura M. Ramzy, Mehabuba Rahman, Manuela Orrego Luque, Kristine Knuti Rodrigues, Robert Belknap, Julie A. Venci, Beatrice Francis, Betsy J. Ruckard, Wendy Moran-Ibarra, Rasulo M. Rasulo, Adrien Matadi, Marisol G. Ramirez, Pa S. Thee, Heather Dawn McFeron, Samantha Pelican Monson, the Tuberculosis Epidemiologic Studies Consortium

## Abstract

**Purpose:** The purpose of this study was to examine the barriers and facilitators experienced by non-U.S.– born persons during the diagnosis and treatment of latent tuberculosis infection (LTBI) in primary care settings, including the impact of culturally and linguistically congruent care navigation.

**Design:** 25 interviews with non-U.S.–born patients, along with focus groups and surveys with 31 primary care team members and leadership, were conducted.

**Setting:** The study was conducted within a network of Federally Qualified Health Center (FQHC) clinics.

**Participants:** Participants were adult non-U.S.–born patients with LTBI and FQHC care team members. A purposefully selected subsample of randomized participants was interviewed.

**Intervention:** Care navigators followed participants randomized to receive care navigation after a positive test for tuberculosis (TB) infection and offered health navigation and education about the importance of TB screening and treatment.

**Method:** Data collection was followed by thematic analysis guided by a critical ideological paradigm.

**Results:** Culturally and linguistically congruent navigation emerged as central to potentially reducing barriers, fostering trust, and improving treatment continuity. Participants without navigation support reported confusion and disengagement from care, while those with culturally aligned navigators described clarity and comfort, with influence overall by intrinsic motivation, relational support, and culturally shaped beliefs about care.

**Conclusion:** Care navigation that includes culturally and linguistically congruent navigators whenever possible may help increase LTBI treatment completion among non-U.S.–born populations. Limitations of the study include the potential influence of cultural norms, power dynamics, and selection bias.

## PURPOSE

Identifying and treating people with latent tuberculosis (TB) infection (LTBI) is a critical component of eliminating tuberculosis in the United States (U.S.).^1^ According to the Centers for Disease Control and Prevention (CDC), progression from untreated LTBI to TB disease accounts for more than 80% of TB cases in the U.S.^2^ Because individuals with LTBI do not exhibit symptoms, they are often unaware of their condition. Without proper treatment, they are at risk of developing TB disease, which is contagious and can lead to significant public health challenges. This makes diagnosis and treatment of LTBI essential for TB elimination in the U.S.^3^

Primary care clinicians are in a key position to identify and treat LTBI, especially in patients who have lived in countries with a higher TB prevalence.^4^ CDC guidelines recommend routine screening for LTBI among individuals born in high TB prevalence countries or those who have had close contact with individuals with TB disease.^2^

Non-U.S.–born patients often face significant barriers to healthcare in the U.S. These barriers are multifactorial, with language differences, lack of familiarity with the U.S. healthcare system, financial limitations, and cultural differences all contributing to the challenges these populations experience when seeking care.^5-9^ Further, the perspectives of these patients are frequently overlooked in healthcare planning, with their experiences and needs less represented in research compared to U.S.-born, English-speaking patients or healthcare clinicians.^10^ Addressing these challenges and including those perspectives is particularly crucial in areas like TB prevention, where non-U.S.–born patients may be at higher risk for LTBI and where a myriad of barriers pose challenges to successful treatment completion. ^11,12^

Previous studies in cancer care have shown that patient navigation, which offers culturally and linguistically congruent care through education, reminders, and cultural translation, can bridge healthcare gaps and reduce differences in health outcomes.^13^ Patient navigators, who are often bi- or multilingual and may have cultural and lived experience congruence with the populations they serve, help patients navigate the healthcare system, and assisting them with scheduling appointments and completing treatment regimens. As Kamaraju et al. (2022) highlight, patient navigation programs have been shown to improve adherence to treatment and follow-up care by addressing the unique needs and concerns of immigrant and refugee patients.^14^

The current project aims include identifying (1) facilitators to diagnosis and treatment of LTBI, (2) barriers to diagnosis and treatment of LTBI, and (3) the role of culturally and linguistically matched care navigation on the LTBI diagnosis and treatment process in primary care settings. The study employs a grounded theory approach, meaning there are no predefined hypotheses guiding the analysis.^15^

## DESIGN

This study employed interviews with non-U.S.–born patients, focus groups with clinic staff members, and surveys administered to clinic leadership. Purposive sampling was used to maximize representation across employee roles and patient regions of origin, intervention groups, and stages in the care cascade (LTBI diagnosis through treatment being declined, ongoing, discontinued or complete).

In 2022, a network of Federally Qualified Health Centers (FQHCs) implemented an electronic medical record (EMR) alert intervention to identify patients eligible for TB screening during primary care visits. EMR alert criteria included ≥2 years of age, birth country with higher TB disease incidence (>20/100,000), and no record of previous screening or treatment for TB or LTBI. Patients with a positive TB screening test result (interferon-gamma release assay) were then randomized to receive care navigation or standard clinic care and followed from the time of a positive TB test through LTBI diagnosis and treatment. Patients were matched with a navigator born in the same World Health Organization region^16^ (culturally congruent) and who spoke the patient’s preferred language (linguistically congruent) when available. Care navigators offered health navigation and education about the importance of TB screening and treatment and addressing barriers to care.

## SETTING

The project took place at Denver Health and Hospital Authority, an academic, safety-net healthcare system in Denver, Colorado within 17 of its FQHC clinics. Denver Health was a member of the Tuberculosis Epidemiologic Studies Consortium (TBESC-III), which aimed to improve LTBI outcomes among non-U.S.–born populations through primary care interventions.^17^

## PARTICIPANTS

Patient participants were aged ≥18 years and diagnosed with LTBI after receiving the EMR alert intervention. We included patients who received care navigation (intervention) and patients who received standard clinic care (control). We included patients completing, not completing, and declining treatment. Of 92 patients approached, 25 consented and completed interviews. Exclusion criteria to this project included age <18, death, relocation, loss to follow-up, or prior LTBI treatment. Staff and leadership participants were employed by the FQHCs and actively involved in the delivery of primary care in the study setting.

## METHODS

The project was approved by Denver Health’s Quality Improvement Review Committee and reviewed by the CDC, which deemed it not research, and conducted consistently with applicable federal law and CDC policy (see 45 C.F.R. part 46. 102 (l)).

For interviews, care navigators contacted eligible participants from a generated list of patients identified as having LTBI and assigned to intervention or control groups. They contacted participants by phone and facilitated electronic informed consent. Interview questions to explore current study aims were developed with internal experts and informed by existing literature (Appendix A). Interviews were conducted by qualitative team members L.R., S.P.M., and M.O.L., who were uninvolved in the LTBI care navigation intervention or in the clinical care of the target patient population. All participants declined a virtual interview including video and chose to complete interviews by phone. Qualified interpreters supported non-English and non-Spanish speakers, and a bilingual research assistant conducted Spanish interviews. Interviews were recorded, transcribed, and checked for accuracy. Interviews occurred between October 2024 and February 2025; they were conducted across a representative number of the World Health Organization regions until thematic saturation was reached, following a critical ideological paradigm.^18^ Participants received a $30 gift card.

Clinic staff focus groups and leadership surveys were designed and conducted with the express purpose of contextualizing patient participant experiences. Staff and leaders across the 17 FQHC clinics were recruited by email and provided electronic informed consent. Three 45-minute semi-structured focus groups (Appendix B) were conducted with staff by S.P.M. in July 2024. They were held outside clinic hours and were recorded and transcribed verbatim. Participants received a $25 gift card. Leadership completed an online survey and were not compensated.

All transcripts and survey responses were de-identified, translated (and back-translated as needed), and imported into Atlas.ti (version 8.0.27.0) for thematic analysis. Co-Investigators S.P.M. and L.R. led and conducted the analysis of patients’ experiences from screening through whatever treatment outcome applied to them at the time of the interview (i.e., declined, discontinued, ongoing, or complete). They developed codes using constant comparison and analytic induction, cataloging key phrases in a codebook.^15^ Research assistant M.O.L. contributed to triangulation to enhance credibility, dependability, and confirmability.^19,20^ Themes and illustrative quotes were finalized collaboratively and member-checked with participants prior to dissemination. Total analysis spanned six months.

## RESULTS

### Sample Characteristics

Twenty five non-U.S.–born patients participated in this study (48% female; 52% male; mean age 48.12 years). See Table 1 for detailed demographic information of patient participants, and Table 2 for information pertaining to care cascade, region of origin, and random assignment to control or care navigation intervention groups.

### Themes and Subthemes

Patterns from patient interview transcripts were consistent with project aims of identifying (1) facilitators to diagnosis and treatment of LTBI, (2) barriers to diagnosis and treatment of LTBI, and (3) the role of culturally and linguistically matched care navigation on the LTBI diagnosis and treatment process, and were organized into five themes supported by illustrative quotes (see Table 3): (1) Motivational Factors (aim one), (2) Supportive Experiences (aim one), (3) Burden and Confusion (aim two), (4) Problematic Systemic Elements (aim two), and (5) Cultural Dimensions (aim three). All participants who discontinued or declined treatment were either in the control group or received culturally and linguistically unmatched congruent navigation (e.g., a Burmese care navigator working with a Spanish-speaking participant). Data collected from staff focus groups and leadership surveys suggested several facilitators from a care team perspective. Specifically, participants stated that when care navigators were involved in care, (1) non-U.S.–born patients were more likely to show for appointments, (2) patients expressed a greater sense of trust and respect for the team, (3) the care team recognized limitations associated with non-U.S.-born patients being new to the culture and to healthcare, and (4) much less miscommunication occurred than when relying on interpretation alone. There was no data available indicating downsides of culturally and linguistically matched navigation raised by staff and leadership.

#### Theme 1: Motivational Factors

Participants expressed a sense of responsibility for their own health and the well-being of others around them, which may have facilitated engagement in LTBI treatment. Personal experiences and a desire to protect family and community members emerged as drivers of treatment uptake. Subthemes included: (a) *Treat now to prevent spread;* (b) *Witness to tuberculosis;* and (c) *Protect family and community*. Overall, participants who completed or were engaged in treatment endorsed *Motivational Factors*, whether they were in the control or intervention group.

*Treat now to prevent spread* described participants emphasized early action to prevent tuberculosis as important, and treatment was considered a proactive and logical measure to avoid future health complications. A participant engaged in treatment stated, *“This tuberculosis, if it comes out, it is easy to spread*… *so if the doctor asks me, I should treat it*.*”* The urgency and seriousness of TB were acknowledged by participants, and early treatment was seen as a sensible strategy to protect long-term well-being.

*Witness to tuberculosis* experienced by family or community members reinforced the perceived seriousness of TB. Participants who had a frame of reference understood the consequences of not treating TB, given that they have witnessed suffering or death. A participant shared, “*I know about tuberculosis*… *a neighbor got sick and I took her to the hospital*… *she had tuberculosis*… *it worries me because I saw how the lady was*.*”* Participants also recalled how TB was neglected or poorly managed in their home countries.

*Protect family and community* described the duty expressed by participants to protect their families and communities. This sense of community responsibility, especially toward vulnerable individuals like children or elderly parents, was a compelling reason to pursue treatment. As stated by a participant, *“I get scared, because for a long time I have been sick with TB what if it will be transferred to the children who are living with me*.*”*

#### Theme 2: Supportive Experiences

Positive interpersonal experiences helped participants feel informed, connected, and supported throughout their LTBI treatment. Subthemes included: (a) *Check-ins and access to results*; (b) *Good care overall by team*; (c) *Trusting relationship with a team member*; and (d) *Thorough explanation*. Overall, participants who completed or were engaged in treatment endorsed *Supportive Experiences*, whether they were in the control or intervention group.

*Check-ins and access to results* included regular follow-ups, timely access to test results, and frequent check-ins through calls, appointments, or the patient portal, all of which helped participants feel supported and informed throughout their LTBI treatment. Check-ins provided reassurance, addressed concerns, and strengthened trust in the care process. As one participant stated, *“they called me very often and they were keeping an eye on things they were very attentive*.*”*

*Good care overall by team* described participants’ experiences with compassionate, flexible, and responsive healthcare teams. When staff were understanding and helpful with scheduling, transportation, or medication access, participants felt supported through their LTBI treatment process. A positive care environment was appreciated, as stated by a participant: *“I have had a very good experience. I have received support from everyone who has taken care of me*.*”*

*Trusting relationship with a team member* illustrated the gratitude expressed by participants for the sense of connection experienced through a close, personal relationship with a healthcare team member. A participant shared, *“The doctor who gave me the medicine treated me like her dad, not a patient*.*”* Shared decision making and respect for participants’ autonomy resulted in participants feeling genuinely cared for as a person, helping treatment feel manageable for participants.

*Thorough explanation*, included care teams taking time to make sure the diagnosis, treatment process, and treatment options were understood, was appreciated by participants. Participants stated that they felt better when receiving clear information about LTBI and instructions regarding what to expect during treatment. As one participant shared, *“I didn’t know if it was active or dormant*… *I became worried, but*… *a doctor personally called me and explained the situation. That made me feel more at ease*.*”*

#### Theme 3: Burden and Confusion

Participants described barriers to LTBI treatment that influenced how they understood, prioritized, and responded to LTBI treatment. Subthemes included: (a) *Existing health conditions;* (b) *Fear of the disease and its implications;* (c) *Confusion about the diagnosis and process;* (d) *Reluctance to take medication*, and (e) *Difficulty accepting treatment for a condition with no symptoms*. Three of these subthemes, included *Existing health conditions, Reluctance to take medication*, and *Confusion about the diagnosis and process* were prevalent among participants in the control group across region of origin and place in care cascade.

*Existing health conditions* include facing multiple, serious health issues alongside LTBI, such as heart disease, diabetes, or recovery from surgery. A participant shared, *“Thank God*… *that hepatitis was gone. And now what are they telling me? I have to undergo another treatment for tuberculosis*.*”* These existing conditions made accepting the diagnosis and tolerating treatment side effects for LTBI, an asymptomatic condition, difficult. Participants expressed that managing other conditions took precedence, leading to delays, interruptions, or refusal of LTBI care.

*Fear of the disease and its implications* included participants’ anxiety and uncertainty about what the diagnosis meant for their health, leading to avoidance, silence, or emotional distress; further, participants stated that they only hear about treatment for those with TB disease in their countries of origin. A participant described this as, *“Well, when I was diagnosed with*… *that disease, I was afraid*… *because you don’t know exactly how it’s going to affect you, when you hear that*.*”* This fear of illness, judgment, and the unknown undermined confidence in the care process.

*Confusion about the diagnosis and process* described participants’ confusion around their LTBI diagnosis or care plan. Inadequate explanations, unclear communication, and hesitation to ask questions left them unsure about what LTBI meant or what to expect, making engagement in treatment more difficult. One participant shared, *“They did explain it to me, but I didn’t understand it very well, and*… *I didn’t want to ask*.*”* Perspectives expressed by staff member focus group participants were consistent, noting observed confusion.

*Reluctance to take medication* illustrated a hesitancy toward taking medications, especially for asymptomatic conditions. Concerns about side effects and preferences for natural recovery were expressed. A participant from The Americas stated, “*I just do not agree*… *with the idea of treating everything with medication or taking medication all the time*.*”* For some, the duration or intensity of side effects related to the LTBI regimen felt burdensome, reinforced reluctance to start or continue treatment. Hesitation to take medications was also noted as a barrier to care during the staff member’s focus groups.

*Difficulty accepting treatment for a condition with no symptoms* described the concept of treating an illness without feeling ill as surprising or unnecessary for participants, making it difficult to accept and/or commit to a treatment for something that did not feel real or urgent. One participant noted that this discrepancy was so striking they wondered if an error had occurred, “*It was very confusing*… *I said, ‘Maybe they got the blood test results mixed up or something*.*’”*

#### Theme 4: Problematic Systemic Elements

Participants reported a paradox in that the healthcare system encouraging LTBI treatment was also a system that presented barriers within its own institutional structure; this theme emerged more frequently among participants who did not receive culturally and linguistically congruent navigation support. Subthemes included: (a) *Cost and inadequate insurance coverage;* (b) *Perspective dismissed;* and (c) *Lack of transparency*. All participants represented in the *Perspective dismissed* subtheme were of African descent and did not have access to culturally matched navigation; they all discontinued or declined treatment. Further, subthemes *Perspectives dismissed* and *Lack of transparency* were more prevalent among participants without culturally and linguistically congruent navigation support.

*Cost and inadequate insurance coverage* includes financial barriers, such as appointment co-pays and uncovered testing and treatment costs, which create significant obstacles. As one participant stated, *“The other[treatment] might be feasible. But that one was not within my financial reach*… *it was not covered by my insurance. So, there is an option, but I can’t have it*.*”* Unlike in some of their countries of origin where testing and treatment were often free or low-cost, participants shared that expenses they faced made initiating or continuing treatment difficult, demonstrating how systemic cost issues undermine access and adherence. Staff member focus group participants echoed this barrier, noting cost as a major barrier to treatment.

*Perspective dismissed* described the experience participants expressed of feeling unheard, minimized, or misunderstood when expressing LTBI-related concerns or sharing their personal history, including prior testing. Participants shared that their LTBI concerns were overlooked by providers who were not directly involved in their LTBI care, as expressed by one participant: *“They keep telling me that’s just your back is hurting, it’s not the TB medicine. But then [another doctor said] ‘the TB medication has reacted in your body and is creating an infection*.*’”* This perception was associated with participants who had lower engagement in care. Staff member focus group participants noted that patients who already completed LTBI treatment and were flagged by the system for TB screening inappropriately seemed frustrated.

*Lack of transparency* illustrated the experiences of participants not feeling fully informed throughout the LTBI diagnosis and treatment process and feeling deceived and misled. Participants were not always made aware of the specific tests being conducted or told why blood tests were being repeated when participants had already undergone the same tests in another country. One participant stated, “…*[the doctor] asked, ‘can I just do all testing?’ I was like, sure that’s fine. All testing is good, but I want to know what it’s for; I didn’t know it was HIV and TB*.*”* This perceived lack of transparency left participants feeling misled once positive results emerged.

#### Theme 5: Cultural Dimensions

Participants’ cultural backgrounds deeply shaped their experiences with LTBI diagnosis and treatment, and influenced how individuals understand, engage with, and respond to their care. Subthemes included: (a) *Communication nuances*; (b) *Faith in God*; (c) *Deference to doctor*; (d) *Relative significance*; and (e) *Intense stigma from misinformation*. Two of the subthemes, *Faith in God* and *Deference to doctor*, were expressed by participants who had completed or were engaged in ongoing treatment. Subthemes *Communication nuances, Faith in God, Deference to doctor*, and *Relative significance* were prevalent among participants without culturally and linguistically congruent navigation support.

*Communication nuances* described how participants shared that even with an interpreter, dialect variations and the use of medical jargon often complicate the understanding of LTBI diagnosis and treatment. A participant stated, *“*…*maybe not even just [help with] translation, because sometimes doctors speak in a way that people*… *have a hard time understanding*.*”* These nuances also included Western ways of communicating not always being understood by participants, such as unfamiliar nonverbal expressions and information conveyed once linearly versus circular pattern with repetition. Data from staff member focus groups and leadership surveys aligned with these sentiments.

*Faith in God* was stated by participants as being a strong, sustained source of resilience and hope throughout their LTBI diagnosis and treatment and helped to cope with uncertainty and challenges. Relying on spirituality for understanding and trusting in a higher power was often intertwined with confidence in medical care. One participant shared, *“as the word of God says, ‘*…*I will get you through all [struggles] in God’s time*.*’ That was what strengthened me*.*”*

*Deference to doctor* described participants’ deep respect for medical providers, which at times led to accepting advice without question. One participant stated, *“I wasn’t afraid because the way my doctor worded it*… *helped me trust that everything would be all right*.*”* Deference also included forgiving offenses or missteps made by medical providers, as viewed through the lens of gratitude for having access to doctors in the U.S. Further, staff member focus group participants shared that some patients appeared to provide the response they think providers want to hear, rather than reporting anything perceived as negative or ungrateful.

*Relative significance* illustrated participants sharing that in comparison to adversities they have been through in their home countries or other hardships they are experiencing now, LTBI is a minor concern. A participant stated, “*some men have [active] tuberculosis, they all have a lot of cough and other things*… *for me [latent tuberculosis] is normal*.*”* In contexts of regional conflict, poverty, or widespread TB infection, LTBI was often seen as “normal” or just one of many health issues, making it less urgent or troubling compared to other life experiences.

*Intense stigma from misinformation* was expressed by participants as a deterrent to engaging in LTBI treatment. Participants shared that LTBI was associated with poor hygiene, immoral behavior, or high contagion risk in their home countries. Misinformation led to participants’ fear and reluctance to disclose their diagnosis to avoid judgment and emotional distress. One participant shared, *“They told me how I could address this with my family, but*… *the professionals will understand me, but my family*… *will not. If I tell this to my husband, he will not understand*.*”*

## CONCLUSION

### Implications of Findings

Non-U.S.–born patients experience unique facilitators and barriers related to the diagnosis and treatment of LTBI, yet the perspectives of these patients are frequently overlooked in healthcare planning and research.^21^ Results align with project aims of identifying (1) facilitators to diagnosis and treatment of LTBI, (2) barriers to diagnosis and treatment of LTBI, and (3) the role of culturally and linguistically matched care navigation on the LTBI diagnosis and treatment process in primary care settings.

The importance of supportive, culturally tailored care emerged from the data as associated with overcoming barriers to LTBI treatment, attending to the first aim of the current study (i.e., “facilitators supporting non-U.S.–born patients”). Specifically, current findings revealed that participants who completed or were engaged in treatment endorsed *Supportive Experiences*, whether they were in the control or intervention group, indicating that perceptions of emotional, informational, and relational support played a critical role in sustaining engagement. This theme aligns with the expectation that individuals who feel supported are likely to continue with treatment.^22^ Additionally, *Motivational Factors*, including (a) knowing someone diagnosed with tuberculosis or (b) having a desire to prevent the spread of the disease and protect family and community members, were commonly expressed by those who had completed or were currently engaged in treatment regardless of intervention or control group. The importance of highlighting these motivational elements when working with non-U.S.–born patients diagnosed with LTBI aligns with previous findings suggesting that motivational factors greatly influence patients’ engagement in LTBI treatment.^23^

The overall themes of *Burden and Confusion* and *Problematic Systemic Elements* fall under the second study aim (i.e., “barriers faced by non-U.S.–born patients”). Data revealed that other health issues, a reluctance to take medication, and perplexity around the process were notable barriers to treatment of LTBI, and a lack of perceived transparency a barrier to diagnosis of LTBI for patients; further, these subthemes were expressed by participants who discontinued or declined treatment or did not receive care navigation. Of particular importance is that all three participants represented in the *Perspective Dismissed* subtheme originated from the African region and discontinued or declined treatment. These findings align with previous studies highlighting the potential benefits of culturally and linguistically congruent navigation alleviating confusion related to logistical burdens and promoting treatment continuation^24^.

Current study findings revealed a protective role of culturally and linguistically congruent navigation in amplifying facilitators of the diagnosis and treatment of LTBI by reducing treatment barriers and mitigating challenges faced by non-U.S.–born patients with accessing treatment for LTBI, addressing the third aim of the current study (i.e., “impact, if any, of culturally and linguistically congruent navigation”). ^22,23^ In particular, three *Burden and Confusion* subthemes and two *Problematic Systemic Elements* subthemes were prevalent among participants without culturally and linguistically congruent navigation support, and all participants who discontinued or declined treatment were either in the control group or received culturally unmatched culturally and linguistically congruent navigation. Staff member focus groups and leadership surveys further support this finding, given that participants stated several examples of culturally and linguistically congruent navigation helping non-U.S.–born patients to better articulate their cultural perspectives and feel understood by their care teams, bridging the cultural gap between patients and providers. The inclusion of these perspectives contextualizes the findings from the non-U.S.–born patient interviews because patients do not necessarily have complete understanding of the framework or normative expectations of the U.S. healthcare system.

Also consistent with the third study aim, *Cultural Dimensions* played a notable role in shaping participants’ healthcare experiences, aligning with previous findings exploring healthcare experiences for non-U.S.–born populations.^25, 26^ For example, Ailinger and colleagues (2010) found similar results demonstrating that deliberately tailoring screening and treatment increases engagement in LTBI care for immigrant patients.^26^ Patient experience is also enhanced when culture is incorporated into treatment (e.g., increased salience of and attention to health information), regardless of impact on outcomes.^27^

### Limitations and Strengths

Limitations of the current study include aspects related to cultural and linguistic differences and inherent power dynamics. First, participants’ responses during interviews may have been influenced by concerns that their feedback could impact care, despite repeated assurances of confidentiality. Cultural norms such as deference to authority, expressions of gratitude like “*Gracias a Dios*,” and reluctance to critique care can all shape narratives in ways that may not fully reflect underlying concerns or unmet needs. Second, participants with positive healthcare experiences, stronger support systems, or comfort engaging in research may have been more likely to agree to be interviewed. The inclusion of eight participants who declined or discontinued treatment helped address this limitation. Third, while interpretation was utilized, miscommunication may still have occurred given the potential for participants’ limited health literacy or unfamiliarity with Western medical frameworks.

Study strengths include the focus on perspective of non-U.S.–born populations often disproportionately excluded from research and practice, providing valuable insight and attending to a gap in the evidence base informing care delivery. Further, representation across region of origin mirroring the non-U.S.–born population at the study site, place in care cascade, and intervention/control groups served as major strength for niche populations, though not generalizable given this is a purposive qualitative sample. The inclusion of languages and dialects not commonly featured in healthcare research allows for nuanced insights into how systems function for non-English speaking patients in the U.S. Finally, the flexibility afforded by use of semi-structured individual interviews enabled a level of depth and richness that adds important qualitative context to existing quantitative data.

## SO WHAT?

### What is already known on this topic?

Non-U.S.–born populations face barriers to LTBI care, and interpretation and conventional care alone often fail to address cultural, linguistic, and systemic challenges, leading to confusion, disengagement, and differences in LTBI outcomes.

### What does this article add?

Five themes emerged: Motivational Factors, Supportive Experiences, Burden and Confusion, Problematic Systemic Elements, and Cultural Dimensions. Culturally and linguistically congruent care navigation in this population was reported by patients to reduce barriers, build trust, and improve treatment continuity. Motivation, relational support, and cultural beliefs shaped engagement and prioritization of care regardless of whether patients received navigation.

### What are the implications for health promotion practice or research?

Interpretation does not replace navigation. Cultural context, personal history, and prior testing should be acknowledged.

## Data Availability

All data produced in the present work are contained in the manuscript

## REFERENCES

1. Williams PM, Pratt RH, Walker WL, Price SF, Stewart RJ, Feng P-JI. Tuberculosis — United States, 2023. Morbidity and Mortality Weekly Report. 3 2024;73:265–270. doi:10.15585/mmwr.mm7312a4

2. Cochran J, Tibbs A, Haptu HH, Paradise RK, Bernardo J, Tierney DB. Scaling Up Latent Tuberculosis Infection Testing and Treatment for Non-US Born Patients in a Federally Qualified Community Health Center. Journal of Immigrant and Minority Health. 12 2023;25:1482–1487. doi:10.1007/s10903-023-01514-0

3. Kim S, Thal R, Szkwarko D. Management of Latent Tuberculosis Infection. Journal of the American Medical Association. 2 2023;329:421. doi:10.1001/jama.2022.24362

4. Tang AS, Mochizuki T, Dong Z, Flood J, Katrak SS. Can Primary Care Drive Tuberculosis Elimination? Increasing Latent Tuberculosis Infection Testing and Treatment Initiation at a Community Health Center with a Large Non-U.S.-born Population. Journal of Immigrant and Minority Health. 8 2023;25:803–815. doi:10.1007/s10903-022-01438-1

5. Gonzalez-Reyes R, Katz D, Lambert L, Sorri Y, Narita M, Horne DJ. Interpreter usage and associations with latent tuberculosis infection treatment acceptance and completion in the USA among non-U.S.–born persons, 2012–2017. PLOS ONE. 4 2024;19:e0298628. doi:10.1371/journal.pone.0298628

6. Dubus N, LeBoeuf HS. A qualitative study of the perceived effectiveness of refugee services among consumers, providers, and interpreters. Transcultural Psychiatry. 10 2019;56:827–844. doi:10.1177/1363461519844360

7. Shaw SA. Bridge Builders: A Qualitative Study Exploring the Experiences of Former Refugees Working as Caseworkers in the United States. Journal of Social Service Research. 5 2014;40:284–296. doi:10.1080/01488376.2014.901276

8. Ho M-J. Sociocultural aspects of tuberculosis: a literature review and a case study of immigrant tuberculosis. Social Science & Medicine. 8 2004;59:753–762. doi:10.1016/j.socscimed.2003.11.033

9. Singh GK, Rodriguez-Lainz A, Kogan MD. Immigrant Health Inequalities in the United States: Use of Eight Major National Data Systems. Scientific World Journal. 1 2013;2013 doi:10.1155/2013/512313

10. Seagle EE, Dam AJ, Shah PP, et al. Research ethics and refugee health: a review of reported considerations and applications in published refugee health literature, 2015-2018. Conflict and Health. 12 2020;14:39. doi:10.1186/s13031-020-00283-z

11. Petersen E, Al-Abri S, Al-Jardani A, et al. Screening for latent tuberculosis in migrants—status quo and future challenges. Int J Infect Dis. 4 2024;141:107002. doi:10.1016/j.ijid.2024.107002

12. Seyedmehdi SM, Jamaati H, Varahram M, et al. Barriers and facilitators of tuberculosis treatment among immigrants: an integrative review. BMC Public Health. 2024/12/18 2024;24(1):3514. doi:10.1186/s12889-024-21020-8

13. Jang J, Sun KH, Mann K, et al. Patient Navigation Increases Breast, Cervical, and Colorectal Cancer Screening Among Immigrants in the U.S.: A Systematic Review. Journal of General Internal Medicine. 7 2025;40:2358–2368. doi:10.1007/s11606-025-09566-8

14. Kamaraju S, Merrill J, Wu J, et al. Patient Navigation Based Care-delivery to Reduce Inequities in Cancer Care Among Immigrants and Refugees: A Commentary on the Successes and the Unmet Needs. International Journal of Cancer Care and Delivery. 3 2022;2 doi:10.53876/001c.33154

15. Strauss A, Corbin J. Basics of qualitative research: Techniques and procedures for developing grounded theory, 2nd ed. Basics of qualitative research: Techniques and procedures for developing grounded theory, 2nd ed. Sage Publications, Inc; 1998:xiii, 312–xiii, 312.

16. World Health Organization. World regions according to the World Health Organization. 2026. https://ourworldindata.org/grapher/who-regions

17. Vonnahme LA, Ravindhran P, Espey J, et al. Latent tuberculosis infection care cascade outcomes in primary care clinics in the tuberculosis epidemiologic studies consortium-III. Annals of Epidemiology. 2026/06/01/2026;118:110081. doi:10.1016/j.annepidem.2026.110081

18. Ponterotto JG. Qualitative research in counseling psychology: A primer on research paradigms and philosophy of science. Journal of Counseling Psychology. 4 2005;52:126–136. doi:10.1037/0022-0167.52.2.126

19. Morrow SL. Quality and trustworthiness in qualitative research in counseling psychology. Journal of Counseling Psychology. 4 2005;52:250–260. doi:10.1037/0022-0167.52.2.250

20. Patton MQ. Evaluation, Knowledge Management, Best Practices, and High Quality Lessons Learned. American Journal of Evaluation. 9 2001;22:329–336. doi:10.1177/109821400102200307

21. Kamran H, Hassan H, Ali MUN, et al. Scoping review: barriers to primary care access experienced by immigrants and refugees in English-speaking countries. Qualitative Research Journal. 7 2022;22:401–414. doi:10.1108/QRJ-02-2022-0028

22. Barik AL, Indarwati R, Sulistiawati S. The role of social support on treatment adherence in TB patients: A systematic review. Nurse and Health: Jurnal Keperawatan. 12 2020;9:201–210. doi:10.36720/nhjk.v9i2.186

23. Heyd A, Heffernan C, Storey K, Wild TC, Long R. Treating latent tuberculosis infection (LTBI) with isoniazid and rifapentine (3HP) in an inner-city population with psychosocial barriers to treatment adherence: A qualitative descriptive study. PLOS Global Public Health. 12 2021;1:e0000017. doi:10.1371/journal.pgph.0000017

24. Shommu NS, Ahmed S, Rumana N, Barron GRS, McBrien KA, Turin TC. What is the scope of improving immigrant and ethnic minority healthcare using community navigators: A systematic scoping review. International Journal for Equity in Health. 12 2016;15:6. doi:10.1186/s12939-016-0298-8

