## Supplemental Materials for "Elevating the patient perspective: Qualitative evaluation of non-U.S.–born care navigation on latent tuberculosis infection screening and treatment adherence"

**APPENDICES**

APPENDIX A: SEMI-STRUCTURED INTERVIEW GUIDE

1. How did you feel about being told you needed to be screened for LTBI?

a. What would have made the experience better?

2. When you were told that you have LTBI and need treatment after your blood test, what were you feeling and thinking?

a. Did you feel like you could ask questions about the diagnosis? If not, what got in the way?

b. What would have made that experience better?

3. Who was most helpful to you throughout the process when you were told you have latent tuberculosis?

a. What specific things did that person help you with?

*i. Prompt if needed: logistics -- transportation, childcare, time off work, helping to find a*	*time that works for them (able to directly schedule), completing treatment?*

*ii. Prompt if needed: relational – feeling understood, could trust them, helped to trust the*	*system more, problem solving, addressing fear, answering questions, appointment*	*reminder calls, etc.*

b. What do you wish they could have helped you with?

4. Did anything about the LTBI treatment process ever feel bad or confusing to you?

a*. Prompt if needed: The way the diagnosis was communicated to you? Feeling “talked down to”? Being asked to come back multiple times for treatment?*

5. Anything else about the LTBI treatment process that you think could make it easier for you?

6. Anything else you’d like me to know or that you’d like to share?

APPENDIX B: SEMI-STRUCTURED FOCUS GROUP GUIDE

1. How is it for you to care for a patient throughout LBTI treatment?

*(listening points/possible probes: What’s difficult? What’s good?)*

2. What factors have you seen have a positive influence on patient adherence to latent TB treatment? *(listening points/possible probes: Language concordance/shared culture between patient and care team member? Addressing stigma? Inclusion of family? Shared decision making? Access to help with transportation? Impact on other conditions and/or healthcare engagement?)*

3. What factors have you seen have a negative influence on patient adherence to latent TB treatment, or that you’ve seen contribute to patients “falling off” or “shutting down”?

*(listening points/possible probes: In response to being told the dx? Pts still holding a lot of stigma around TB? When they have to return for xrays, PharmD appointments, etc? other points of confusion, myths?)*

4. What do you think would help address any patient-related issues you’ve noticed, or make it easier for immigrant and refugee patients?

*(What do you think would help keep patients engaged in a conversation about LTBI? Having family involved in the process? Always using an interpreter that’s a non-family member? Adding incentives? Ensuring access to transportation and other resources?)*

5. What particular person or role in the clinic is most helpful in engaging patients? Like who do you see patients calling or asking for by name?

*(listening points/possible probes: Have you had a care navigator involved in LTBI patient care in the clinic?)*

6. Anything else you’d like me to know or that you’d like to share about patients and LTBI?

TABLES

Table 1. Characteristics of interview participants diagnosed with latent tuberculosis infection while undergoing TB evaluation (N =25): 2024-2025

| Characteristic | N | **%** |
| --- | --- | --- |
| **Sex at birth** |  |  |
| Male | 13 | 52% |
| Female | 12 | 48% |
| **Age, years** |  |  |
| 18-29 | 2 | 8% |
| 30-39 | 6 | 24% |
| 40-49 | 5 | 20% |
| 50-59 | 6 | 24% |
| 60-69 | 6 | 24% |
| **Region of Origin*** |  |  |
| The Americas | 13 | 52% |
| Eastern Mediterranean | 5 | 20% |
| Africa | 5 | 20% |
| Southeast Asia | 2 | 8% |
| **Insurance Status** |  |  |
| Uninsured | 12 | 48% |
| Medicaid | 8 | 32% |
| Private Insurance | 3 | 12% |
| Medicare | 2 | 8% |
| **Language of Origin** |  |  |
| Spanish | 13 | 52% |
| Amharic | 2 | 8% |
| Dari | 2 | 8% |
| Arabic | 2 | 8% |
| Vietnamese | 2 | 8% |
| Pashto | 1 | 4% |
| Farsi | 1 | 4% |
| English | 1 | 4% |
| Oromo | 1 | 4% |
| **Preferred Language**** |  |  |
| Spanish | 13 | 52% |
| English | 5 | 20% |
| Amharic | 2 | 8% |
| Vietnamese | 2 | 8% |
| Dari | 1 | 4% |
| Arabic | 1 | 4% |
| Oromo | 1 | 4% |

**Based on World Health Organization categorization*

***Language preferred to use during interview*

Table 2. Treatment outcomes among people diagnosed with latent tuberculosis infection by intervention group, cultural and linguistic concordance of care navigator, and world region of origin (N=25): 2024-2025

|  | **The Americas** | **Africa** | **Eastern Mediterranean** | **Southeast Asia** |
| --- | --- | --- | --- | --- |
| **Completed Treatment** | Intervention* Intervention* Intervention* Intervention* | Control^+^  Control | Intervention^+^  Intervention^+^ |  |
| **Ongoing Treatment** | Intervention*  Intervention*  Control  Control  Control | Intervention* | Control  Control^+^ | Intervention |
| **Treatment Discontinued** | Intervention  Intervention  Intervention | Control |  | Control |
| **Declined Treatment** | Control | Control | Control |  |

*^* = Care Navigator and participant were culturally and linguistically matched in that they were born in the same world region and spoke the same native language^*

*^+ = English speaking participant^*

*^Note: Intervention = received care navigation; Control = received care as usual^*

Table 3. Illustrative participant quotes reflecting themes influencing LTBI treatment decisions, by world region of origin, treatment status, and care navigation allocation (N=25): 2024–2025

Note: Interviewer initials precede participant number.

| **MOTIVATIONAL FACTORS** | |
| --- | --- |
| ***Treat Now To Prevent Spread*** | "Yes, if it is possible to prevent things from happening, why not prevent them? Why leave them?"  *~MOL06 (The Americas, Completed, Intervention)* |
|  | "Focusing on [prevention] in your first appointment, or even before that, is important... approaching the people to educate and prevent."  *~MOL08 (The Americas, Ongoing, Control)* |
|  | "The magnitude of the disease and the situation—I'm well aware of it. If you already have a diagnosis, no matter what, you have to receive treatment because I know it can get complicated later. Maybe not now, but later on, over time, it can affect my health. So, I think... it is better to get treatment right away."  *~MOL01 (The Americas, Ongoing, Control)* |
| ***Witness to Tuberculosis*** | "When the doctor said, 'Your tuberculosis is inactive,' it was good news for me. I was trying to tell the doctor, 'Please give me some medicines to use it in order to save me because one of my brothers became disabled because of TB in [the Eastern Mediterranean region].' He's younger than me. Unfortunately, now he cannot walk."  *~LMR03 (Eastern Mediterranean, Completed, Intervention)* |
|  | "Seven or eight years ago [my uncle] passed away, and he had it."  *~LMR11 (Eastern Mediterranean, Ongoing, Control)* |
|  | "I had a sister-in-law in my country who got active tuberculosis because, as I mentioned, in my country they don’t do these tests when it’s dormant. They wait until you’re already dying."  *~MOL07 (The Americas, Ongoing, Control)* |
| ***Protect Family and Community*** | "If this disease is dormant, if it is latent, it can reactivate at any moment... If this thing reactivates and I don’t receive that treatment... I feel kind of guilty as another person is going to be hurt because I didn't want to have this treatment."  *~MOL09 (The Americas, Ongoing, Control)* |
|  | "I'm old. What is the fact that you accept to let people treat your latent tuberculosis? What is the factor that most affects that decision? There are small children in my family. If I don't treat it, it will affect young children, so I agree to treat."  *~SPM03 (Southeast Asia, Ongoing, Intervention)* |
|  | "We could diagnose it and then have the treatment, and I will not make another person sick. So, I'm really happy for myself, and also for the community. I'm happy for my family... So that's a good thing."  *~LMR11 (Eastern Mediterranean, Ongoing, Control)* |
| **SUPPORTIVE EXPERIENCES** | |
| ***Check-ins and Access to Results*** | "I have the app...they send me everything there. As soon as they complete the tests, they send them to me there or email them to me. And I look at the app and check my email, and I see everything. Then someone contacts me."  *~MOL06 (The Americas, Completed, Intervention)* |
|  | "The pharmacist was great. She explained to me all the treatment, and we had a phone call, and I think I was seeing her once a month."  *~LMR06 (Africa, Completed, Control)* |
|  | "Yeah, they were calling me after a month; when they were calling me, [they ask], 'you had any problem with your medication?', and it was good for me."  *~LMR03 (Eastern Mediterranean, Completed, Intervention)* |
| ***Good Care Overall by Team*** | "I had a great experience with the team of doctors, nurses and all the workers. They're the ones who have taken care of me very well. Sometimes I arrive a little late and I tell them, 'The bus was late,' I’m maybe 10 or 15 minutes late. 'No, don’t worry, you are going to come in,' they say. They give [me] understanding and flexibility." *~MOL04 (The Americas, Ongoing, Control)* |
|  | "It was a great clinic, and it, they did help me a lot. Everything was clear, so I didn't have any questions." *~LMR01 (Africa, Ongoing, Intervention)* |
|  | "It seems [my prescriptions] weren't ready yet, but the guy told me, 'We don't have the prescription you ordered yet; it's not here yet.' But then he told me, 'But I'm going to do it now, in 10 minutes.' And I sat in the chair in the clinic. And in 10 minutes, my name appeared on the screen, and they gave me the medicines quickly." *~MOL09 (The Americas, Ongoing, Control)* |
| ***Trusting Relationship with a Team Member*** | "She is one of the best nurses, and she's a really good nurse. ...and I'm always with her, I always have an appointment with her. And so I'm happy that we started [treatment]. *~LMR03 (Eastern Mediterranean, Completed, Control)* |
|  | "The doctor who gave me the medicine treated me like her dad, not a patient." *~LMR07 (Eastern Mediterranean, Completed, Intervention)* |
|  | "Yes, the pharmacist, the one who prescribed the medication, she explained the symptoms I would experience and the changes in my body. She also told me to call if anything happened, and they gave me her number. " *~MOL07 (The Americas, Ongoing, Control)* |
| ***Thorough Explanation*** | "She explained to me, 'It's like it's TB, but it's sleeping...stuff like that. But there is a chance it will wake. So she explained it very well."  *~LMR06 (Africa, Completed, Control)* |
|  | "Yes, I talked to the doctor. I asked her about the process. I asked her why I had gotten sick. I also asked her what the side effects might be during the course of my treatment and all that. And she explained it all to me."  *~MOL05 (The Americas, Ongoing, Intervention)* |
|  | "I liked everything. The way she explained everything, giving me information beforehand.... And she told me everything in detail. She explained the situation well, and what I had to do." *~MOL03 (The Americas, Ongoing, Intervention)* |
| **BURDEN AND CONFUSION** | |
| ***Existing Health Conditions*** | "I have a stent, and I've just had another stent implanted. So, at that time, I was taking a lot of medications. And taking those medications plus [LTBI treatment], I couldn't stay awake. That was the reason why I had to suspend the treatment. There was no other option for me." *~MOL10 (The Americas, Discontinued, Intervention)* |
|  | "I had an ear surgery. My blood pressure went down when I took the medication for that surgery. I was there for 2 hours, in the ED. Then [LTBI treatment] felt like too much, not something I wanted to deal with." *~LMR04 (Eastern Mediterranean, Declined, Control)* |
|  | "I'm a diabetic, right? So right there, I got really bad with my diabetes. I got a renal virus. Then I got complications in my lungs, and they had to put oxygen on me for 2 to 3 months. And then that's when they said, 'Hey, you have latent tuberculosis.'" *~LMR12 (The Americas, Completed, Intervention)* |
| ***Fear of the Disease and its Implications*** | "It's fear of what it does to you...it's fear of infecting other people, fear that what you have, let's say, that others judge you. It's fear of not understanding what they're telling you." *~MOL09 (The Americas, Ongoing, Control)* |
|  | "Yes, I was scared; how would the medications affect me, that is, emotionally, how would they affect other organs? ...What is that going to be like?"  ~*MOL05 (The Americas, Ongoing, Intervention)* |
|  | "I don't tell anyone anything... it scared me more, they made me more nervous. So, I preferred to keep it to myself."  *~MOL11 (The Americas, Discontinued, Intervention)* |
| ***Confusion About the Diagnosis and Process*** | "So by the end of these three months [of LTBI treatment], one big [question], do I still have [the bacteria] in my body? Or if it will be gone?"  *~LMR09 (Africa, Ongoing, Control)* |
|  | "The process was a bit fractioned. I mean, at the beginning, they told you, 'oh, you have latent tuberculosis.' I had many questions in my head—many questions and doubts.  ... And then the next step, the next person didn't know what to tell you, so the next person treated you. But it was for something else, so there was no communication or anything."  *~MOL01 (The Americas, Discontinued, Control)* |
|  | “I was kind of confused because I was told that I have a different kind of problem. But when he mentioned the results, I was kind of confused. Yeah, that’s part of why I’m asking these questions, because it’s really confusing.  ~ *LMR10 (Africa, Declined, Control)* |
| ***Reluctance to Take Medication*** | "It has not been easy. Sometimes I feel a reaction to the medication. I feel a little tired...a little bit allergic, but the doctor told me this was going to happen." *~MOL04 (The Americas, Ongoing, Control)* |
|  | "I personally don’t take medications unless I’m really sick or I need it, like now.  When I am sick with the flu or a cold or something...I try to get better naturally. I do not like to take many pills." *~MOL06 (The Americas, Completed, Intervention)* |
| ***Difficulty Accepting Treatment for a Condition with No Symptoms*** | "It doesn't make sense to take medicine because I don’t feel sick, it’s not active."  *~LMR04 (Eastern Mediterranean, Declined, Control)* |
|  | "I didn’t feel anything, not even a headache, no body aches, no tiredness, no sweating, nothing. I told her I didn’t feel anything. At all."  *~MOL08 (The Americas, Ongoing, Control)* |
|  | "When the doctor gave me the tuberculosis diagnosis, I said, ''Wow, they made a mistake," because I thought to myself, 'I don't have any symptoms. I feel fine.' I was really puzzled because the word tuberculosis always causes a lot of fear in people. "  *~MOL01 (The Americas, Ongoing, Control)* |
| **PROBLEMATIC SYSTEMIC ELEMENTS** | |
| ***Cost and Inadequate Insurance Coverage*** | "In [the Eastern Mediterranean region], which is a poor country, it was free for everyone. [In the US], they tell me that every appointment, you have to pay $15. I paid that $15. Before the next appointment, they sent me a $500 bill. Then I was scared, and I didn't go to the next appointment. I just cancelled my appointment, and I didn't take any other appointments."  *~LMR03 (Eastern Mediterranean, Completed, Intervention)* |
|  | "I stopped taking the medication [due to side effects]. And that other treatment, which could be feasible, is expensive and not covered by my insurance. I don't have the financial means to pay for it. So, that's it. That's how I ran out of options. I fought for it as much as I could, but no more."  *~MOL10 (The Americas, Discontinued, Intervention)* |
|  | "The questions were always in my head. It always made me very uneasy. I knew this disease could be treated, but I was also very worried about the cost of the treatment and the time it would take."  *~MOL01 (The Americas, Ongoing, Control)* |
| ***Perspective Dismissed*** | "So whenever I go to the clinic, my son or my daughter talk on behalf because I don't speak English. Okay, but I still feel like that's not right because you should also be able to participate in the conversation about your body and your health, right?"  ~*LMR10 (Africa, Declined, Control)* |
|  | " And at the end... I was thinking, okay, I'm gonna be done with this, and it's gonna be, you know, out of my body. And then they told me, no, there's actually still a very small chance that it could... So why am I putting a bunch of medicine in my body? I said I don't like taking medication."  *~LMR06 (Africa, Completed, Control)* |
|  | "I've been consistently telling them that I've been having the worst [side effects], and I've been begging every day, 'Should I stop the medication?' They say, 'No, no, no!' And as they say, 'No,' [the side effects] keep worsening, and it almost took my life."  *~SPM02 (Africa, Discontinued, Control)* |
| ***Lack of Transparency*** | "They didn’t ask specifically if they can test for latent TB, just did the xray. I wish they would have asked."  *~LMR08 (Southeast Asia, Ongoing, Control)* |
|  | "Well, um, it was a general test, I didn’t... They never explained it was a tuberculosis test. They just said it was a general test."  *~MOL03 (The Americas, , Ongoing, Intervention)* |
|  | "They didn't tell me exactly what they were testing. They didn't tell me about the lung testing. And if they did, I didn't know."  *~LMR10 (Africa, Declined, Control)* |
| **CULTURAL DIMENSIONS** | |
| ***Communication Nuances*** | "If I get confused about something, [I need] someone coming over and helping me with the translation. And maybe not even translation, but sometimes doctors speak in a way that people who are not working in healthcare have a hard time understanding."  *~LMR08 (Southeast Asia, Ongoing, Control)* |
|  | "Farsi is a dialect that Iranian people [speak]. It's the same language [as what Afghan people speak], but a different dialect. So sometimes it's difficult to have a good communication with someone who speaks Farsi instead of Dari."  *~LMR11 (Eastern Mediterranean, Ongoing, Control)* |
|  | "Having someone who speaks your language explaining the treatment and giving you the diagnosis is useful. Of course."  *~MOL11 (The Americas, Discontinued, Intervention)* |
| ***Faith in God*** | "I have held onto God and told Him, 'Lord, you are my ultimate doctor.' He’ll take control of everything."  *~MOL04 (The Americas, Ongoing, Control)* |
|  | "I don't know what to say, but I only trust God. Everything that happened was just part of destiny."  *~SPM02 (Africa, Discontinued, Control)* |
|  | "I trust my doctor, but my faith is placed in God."  *~MOL08 (The Americas, Ongoing, Control)* |
| ***Deference to Doctor*** | "I said, 'Well okay, I have to have those tests because if the doctor is telling me to have them, it's for a reason.' I thought, 'There’s a reason for it. Maybe he saw something. He hasn't told me anything, but he might have seen something and wants to make sure that he can tell me something."  *~MOL09 (The Americas, Ongoing, Control)* |
|  | "Maybe she was busy.... but this is how it felt. I'm sure she didn't mean to [offend me]... But you know, when you go home and you think about it, there's always those afterthoughts."  *~LMR06 (Africa, Completed, Control)* |
|  | "I believe in doctors so I said, 'This medication will be fine because, of course, it’s already possible to prescribe it to patients.'"  *~MOL02 (The Americas, Ongoing, Intervention)* |
| ***Relative Significance*** | "I'm tired of hearing that I have all these problems [including LTBI]. In [the Eastern Mediterranean region], having these issues is normal. It's not a big deal. ...50% of people have it. We’re just trying to survive [terrorist group]." *~LMR04 (Eastern Mediterranean, Declined, Control)* |
|  | "Unfortunately, as we come from Africa cultures and Africa countries, we're used to [living with LTBI] now. There's no choice, and you get on with it."  *~MOL10 (The Americas, Discontinued, Intervention)* |
|  | "I didn't ask anybody to come with me. This is not something big like, not a big deal."  *~LMR09 (Africa, Completed, Control)* |
| ***Intense Stigma from Misinformation*** | "I also felt kind of sad, because I have heard people around me say this disease is common among people who don’t have good hygiene.... and I am really not like that. Actually, I've kept it very discreet because... if I say, 'I have this,' they're going say, 'Oh, she's got that! I'm not getting close to her.'"  *~MOL05 (The Americas, Ongoing, Intervention)* |
|  | "My partner knows about my diagnosis, but he still talks it as if it weren't true; 'You don't have that.' He's not really supportive because he says, 'No, don't mention that. No, don't talk too much about that disease. It's very ugly.'  *~MOL01 (The Americas, Ongoing, Control)* |
|  | "I don't tell anyone anything, just my family and my mom. But after [LTBI treatment], I didn't even tell them anymore because they scared me more, they made me more nervous. So, I prefer to keep it to myself."  *~MOL11 (The Americas, Discontinued, Intervention)* |

Table 4. Implications for practice from a Federally Qualified Health Center system serving non-U.S.–born patients with latent tuberculosis infection: 2024–2025

| Theme | Implication | |
| --- | --- | --- |
| Theme 5: Cultural Dimensions;  subtheme “*Communication nuances*”  Theme 3: Burden and Confusion;  Subtheme  *“Confusion about the diagnosis and process”* | **Do Not Assume Interpretation is the Same as Care Navigation** | Language interpretation is not interchangeable with care navigation. Culturally congruent care navigators can shift from linear, transactional dialogue to relational approaches better aligned with non-U.S.–born patients’ linguistic norms, and can recognize culturally specific expressions of emotion that are not typically conveyed through interpretation alone.  As such, care navigation may complement interpretation in delivering supportive, culturally responsive care. |
| Theme 1: Motivational Factors  Theme 3: Burden and Confusion  “*Difficulty accepting treatment for a condition with no symptoms”*  Theme 5: Cultural Dimensions;  Subthemes *“Relative significance”* | **Keep Cultural Context in Mind** | To support adherence, care teams should identify and leverage culturally and contextually meaningful motivators, including faith, stigma, and the significance of LTBI. Framing treatment as protecting family and community may resonate more than abstract health benefits. Staying attuned to patients’ lived experiences and ensuring they feel heard, respected, and validated is essential to building trust.  Providers and systems should recognize that agreement with care recommendations—especially in the asymptomatic LTBI care cascade—may not reflect true intention to follow-through. Patients may appear compliant during encounters while competing life demands, such as socioeconomic instability, complex family responsibilities, continuous acculturation, or transcontinental living, take precedence. |
| Theme 4: Problematic Systemic Elements;  Subtheme *“Perspective dismissed”*  Theme 2: Supportive Experiences;  Subtheme  *“Thorough explanation”* | **Recognize and Acknowledge Prior Testing** | It is important to note that many patients may have undergone LTBI testing—and possibly treatment—prior to their current interaction with the healthcare system. Repeated requests for testing across different systems can feel intrusive, particularly for immigrant and refugee populations who may already feel singled out.    To address this, clinicians and care teams should proactively name and normalize the repetition. Clear, empathetic communication that contextualizes testing as part of a public health approach—and reinforces that effective treatment is available—can help support informed decision-making. |
| Theme 2: Supportive Experiences;  Subtheme “Trusting relationship with a team member”  Focus Group and Leadership Survey Findings | **Explore a Centralized Care Navigation Infrastructure** | Future efforts within FQHCs or other medical homes that serve non-U.S.–born patients should explore the feasibility of a centralized “navigation bank” to support culturally and linguistically concordant care across health systems. In systems like the FQHC setting in which this study was conducted, where more than 50 languages are routinely spoken, achieving language and cultural alignment at the point of care is a persistent challenge. A centralized model could allow trained navigators to serve multiple regions or systems, improving scalability and addressing workforce limitations while also prioritizing the relational continuity needed to build trust and maintain engagement.  A centralized model could be leveraged beyond LTBI care to address other pressing public health priorities—such as chronic disease management, vaccine uptake, maternal health, and HIV care—particularly in culturally and linguistically diverse populations where tailored support is most needed. |
